# Covid-19 Excess Mortality in China: A Regional Comparison

**DOI:** 10.1101/2023.06.15.23291443

**Authors:** Lee Liu

**Affiliations:** University of Central Missouri, Warrensburg, MO 64093, USA

**Keywords:** Covid-19 excess mortality, excess death rate, regional disparities, health disparities, China

## Abstract

Estimates of Covid-19 excess mortality are often considered to reflect the true death toll of the pandemic. As such, information on excess mortality is urgently needed to better understand the impact of the pandemic and prepare for future crises. This study estimated Covid-19 excess mortality at the provincial, regional, and national levels in China and investigated its associated regional disparities. The analyses were based on population and death rates data published by the national and provincial bureaus of statistics in China. The results suggest that excess deaths in China were over 1 million during each year of the pandemic, totaling to over 4 million by the end of 2022, at an excess death rate of 15.4%. This rate was likely comparable to that of the Organization for Economic Cooperation and Development (OECD), but higher than the US rate. Striking disparities were discovered among the 31 provinces with excess death rates ranging from negative rates in two eastern provinces to over 30% in three inland provinces. Rates in western China were over twice as high as those in eastern China. Variations with each individual regions were the largest in the central region and the smallest in the Northeast, which was the hardest hit with excess death rate of over 23%. The regional disparities in excess mortality rates seem to reflect pre-existing regional inequalities in socio-economic development in China. Such findings suggest that China has far to go to mitigate regional inequalities, achieve sustainability, and prepare for the next major crises.

## Introduction

The world is urgently in need of further information on Covid-19 excess mortality for a comprehensive understanding of the pandemic’s impacts and better preparation for the next crisis (OECD, 2023; Covid Crisis Group, 2023; WHO, 2023; Sachs et al., 2022). Indeed, excess mortality reflects the true death toll of the Covid-19 pandemic (WHO, 2023). Such information is particularly lacking on China, a country with a population of over 1.4 billion, where the pandemic first started. Initially, the outbreak was concentrated in Wuhan and the surrounding cities in Hubei province, leading to strict lockdown measures and a significant strain on the local healthcare system (Liu, 2020). As the virus spread to other parts of China, the central government implemented various containment measures, including travel restrictions, quarantine measures, and lockdowns. Regional inequalities in healthcare infrastructure and resources became apparent during the pandemic (Liu, 2020). Developed regions in eastern China, such as Beijing, Shanghai, and Guangdong, had better-equipped healthcare systems to deal with the crisis compared to less-developed regions in other parts of China. This disparity resulted in challenges for underprivileged regions in providing adequate medical care, testing, and treatment for their populations.

This study aimed to estimate China’s Covid-19 excess mortality at the provincial, regional, and national levels and explore regional disparities in excess mortality. China is a vast country with significant variations in economic development, infrastructure, and social services between different regions (Veeck et al., 2021). Regional inequality in China has been a long-standing issue and there is a large literature on the topic (eg. Xie and Zhou, 2014). China has taken a series of measures to reduce such inequalities and implement the UN 2030 Agenda with Sustainable Development Goals (SDGs) that require equality and inclusion in developing sustainable communities (UN, 2015). These measures included the Western Development Strategy, the “Go West” campaign, poverty alleviation, regional integration, environmental sustainability, and improving access to quality education and healthcare services in underdeveloped regions. Important progress has been reported (Wang et al, 2022). However, the pandemic can be seen as a test on China’s achievement in reducing regional disparities. Regional inequalities in socio-economic development are likely to have manifested in disparities in Covid-19 excess deaths among the 31 provinces and four regions in China. Recent studies have indicated that many of the challenges during the Covid-19 pandemic were closely related to sustainability issues. In fact, more sustainable US states and OECD countries tended to have better Covid-19 outcomes (Liu, 2023a, 2023b). To place China’s excess mortality in an international context, the 31 provinces will be compared with the 50 US states and the 35 countries in the Organization for Economic Cooperation and Development (OECD).

### Terminologies, Data, and Methods

The 31 provincial-level regions in mainland China include 22 provinces, 4 municipalities (Beijing, Tianjin, Shanghai, and Chongqing), and 5 autonomous regions (Inner Mongolia, Guangxi, Tibet, Ningxia, and Xinjiang). Generally, this study refers to all of them as provinces, and mainland China as China. China officially places the 31 provinces into four regions (NSB, 2023a). The Eastern region comprises of the 10 eastern coast provinces. The remaining 21 provinces include six in the central region, 12 in the western region, and three in the Northeast.

Excess deaths refer to the number of deaths that exceed the expected number of deaths based on historical data. It is used as an indicator of the overall impact of a particular event or condition, such as a pandemic or natural disaster (CDC, 2023). Estimating excess deaths can be done using different methodologies and the results may vary accordingly. The US CDC presents weekly estimates of excess deaths in the US states calculated by Farrington surveillance algorithms (CDC, 2023). The OECD measured excess deaths as the percentage changes in annual all-cause mortality between 2015–2019 and 2020–2021 (OECD, 2023). The methodology used by the OECD was adopted for this research, using the formula:

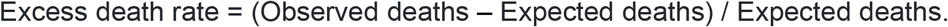

The study calculated provincial excess deaths and death rates based on population and death rates data from 2015 to 2022. The averages of the 2015–2019 population and death rates were used to derive the baseline of population, deaths, and death rates for the 31 provinces. The baseline data were compared to the 2020, 2021, and 2022 data to derive all-cause excess deaths and death rates. The 2015 to 2021 data were from China statistical yearbooks published by the National Bureau of Statistics of China (NBS, 2023a), except for the 2020 death rates which were excluded from the 2021 yearbook. The 2020 death rate and 2022 population and death rate data were gathered from provincial level Economic and Social Development Statistical Bulletins through the provincial bureaus of statistics whose web links were provided by the NBS (NBS, 2023b). The 2022 provincial population data were verified with reports by the media such as China News Service (2023). Xingjiang’s 2020 death rate and Tianjin’s 2022 death rate were unavailable, and their 2021 death rates were used as substitutes.

Using the SPSS software, independent samples t-tests were conducted to compare the means between different Chinese regions to explore regional disparities in China. Additionally, the 31 provinces were compared with the 50 US states and the 35 countries in the Organization for Economic Cooperation and Development (OECD).

## Results

A summary of the main discoveries is as follows:

1. The results indicate large differences in excess all-cause mortality rates among the 31 provinces (Table 1). The highest rates (over 30%) in 2020–2022 were found in Anhui, Xinjiang, and Shanxi (Figure 1). Anhui had the worst rate in 2020 and 2021 while Shanxi’s rate of 38.45% was the highest in 2022. On the other hand, Jiangsu and Beijing stood out as provinces with the lowest rates. Jiangsu experienced negative excess deaths in 2020 and 2021, resulting in an excess death rate of –2.72, meaning fewer annual deaths in the 2020–2022 period than in the base years of 2015–2019. This was a striking difference when compared to its neighbor, Anhui. The least and most impacted provinces were next to each other. The next lowest rate was 2.22% in Beijing, which had a negative rate in 2020. The negative rates accounted for 50,750 fewer deaths during the three years in Jiangsu and Beijing.
2. Hubei’s 2020 rate of 10.25% was lower than might have been expected, considering that was where Wuhan, the ground zero of the Covid-19 pandemic, was located. Being sealed off from the rest of China for months, Hubei might have been feared as the worst impacted province in 2020. However, its rate was lower than its neighboring provinces of Shaanxi and Hunan and was less than one third of its neighbor Anhui’s 34.69%. Hubei’s three-year rate was 14.38%, still below the national rate of 15.4%.
3. China’s excess deaths were over 1 million each year, and over 4 million by the end of 2022 (Table 2). The number of excess deaths increased from 2020 to 2022 in all regions of China. The largest increase was over 88% in the Eastern region. Nationwide there was an over 50% increase. The regional disparities were mainly between the East and the rest of China, as variations were small among Central, Western, and Northeast China. This was true every year. Eastern region’s 2020–2022 rate of 9.07% was less than half of that in the rest of China. The Northeast was the hardest hit region with excess death rate over 2.5 times as high as that in Eastern China.
4. Variations within the regions were also large. Zhejiang had an above national rate, although it is in the Eastern region. As indicated by the large standard deviation values, variations were the largest in the Central region which included two of the worst impacted provinces, Anhui and Shanxi, and some provinces such as Henan that was above the national average (Figure 1). Excluding these two provinces, the Central region would have had an excess death rate of 11.46%, which is not far above the rate for eastern China. The Western region was not entirely bad either. It had three provinces with rates lower than the national average (Guizhou, Chongqing, and Qinghai). Four of the five autonomous regions (Xinjiang, Inner Mongolia, Ningxia, and Tibet) had a rate above the national level, while Guangxi’s rate was below the national rate. The 2020–2022 excess death rate for all five autonomous regions was 21.49%.
5. The independent samples t-tests statistically confirmed a significant difference in excess death rates between Eastern China and the rest of China, as indicated by the t and p values (Table 3). The results were consistent for the whole 2020–2022 period. The effect sizes, as measured by Cohen’s d, were all above 0.9, indicating a large difference between the regional means, with the largest between Eastern and Northeast regions.
6. For an earlier report, the author calculated the excess death rates for the 50 US states based on US CDC’s weekly excess deaths reports and found that the rate was 11.7% by the end of 2021 (Liu, 2023a). Independent samples t-tests revealed that the US rate was significantly lower than the Chinese rate (13.75%) (Table 4). The Cohen’s d of 0.632 indicated a noticeable difference between the 31 provinces and the 50 states. On the other hand, the Chinese provinces were not statistically different from the OECD countries in terms of excess death rates in 2020–2021. The rate was 13.6% among OECD’s 35 member countries (OECD, 2023). The standard deviation values indicated that disparities were smaller among the 31 provinces than among the 35 OECD countries, but higher than among the 50 US states where excess death rates ranged from 2.13 in Hawaii to 18.19 in Arizona (Liu, 2023a).

**Table 1.**
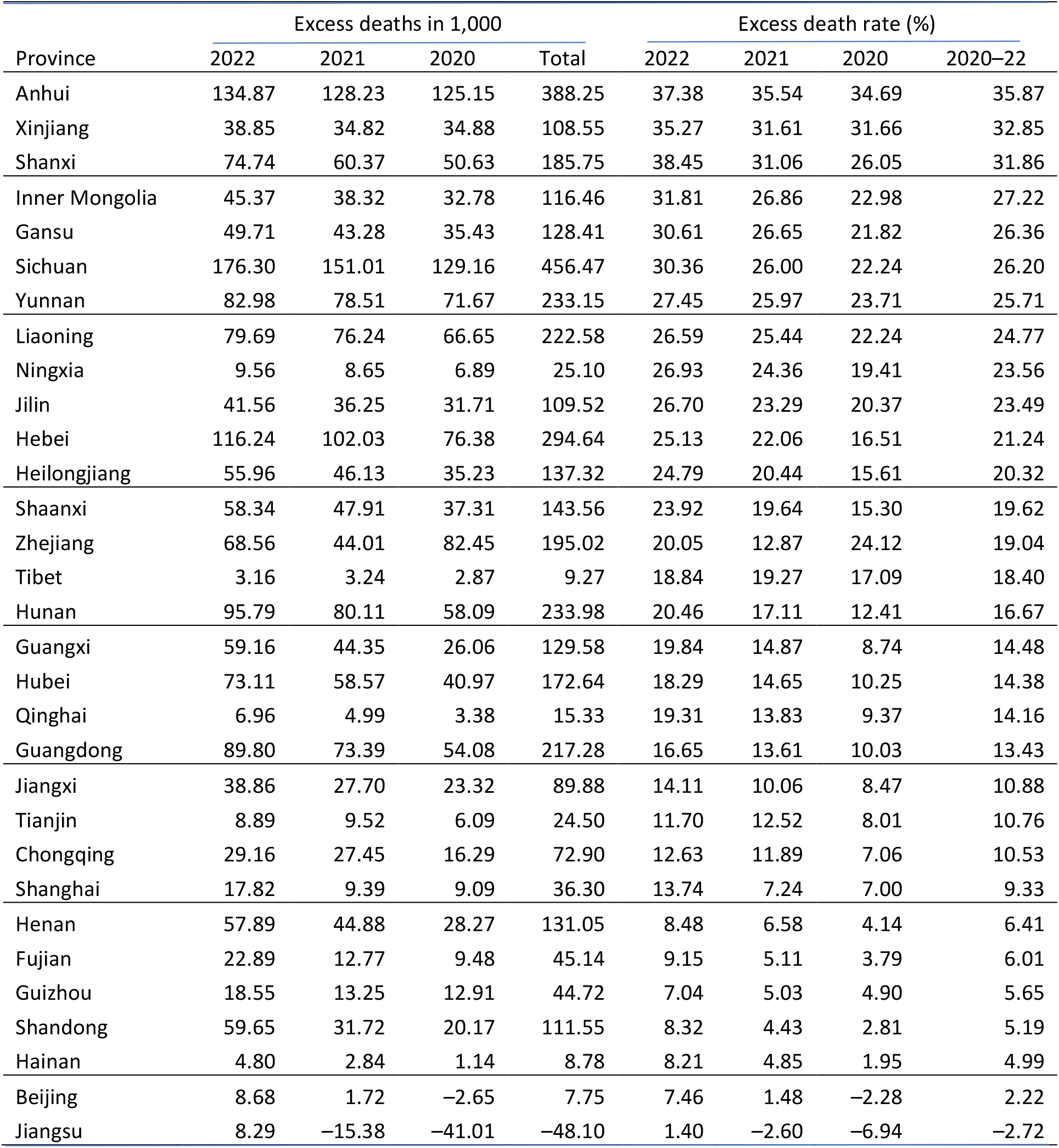
Excess deaths and death rates among the 31 provinces, China, 2020–2022.

**Figure 1.**
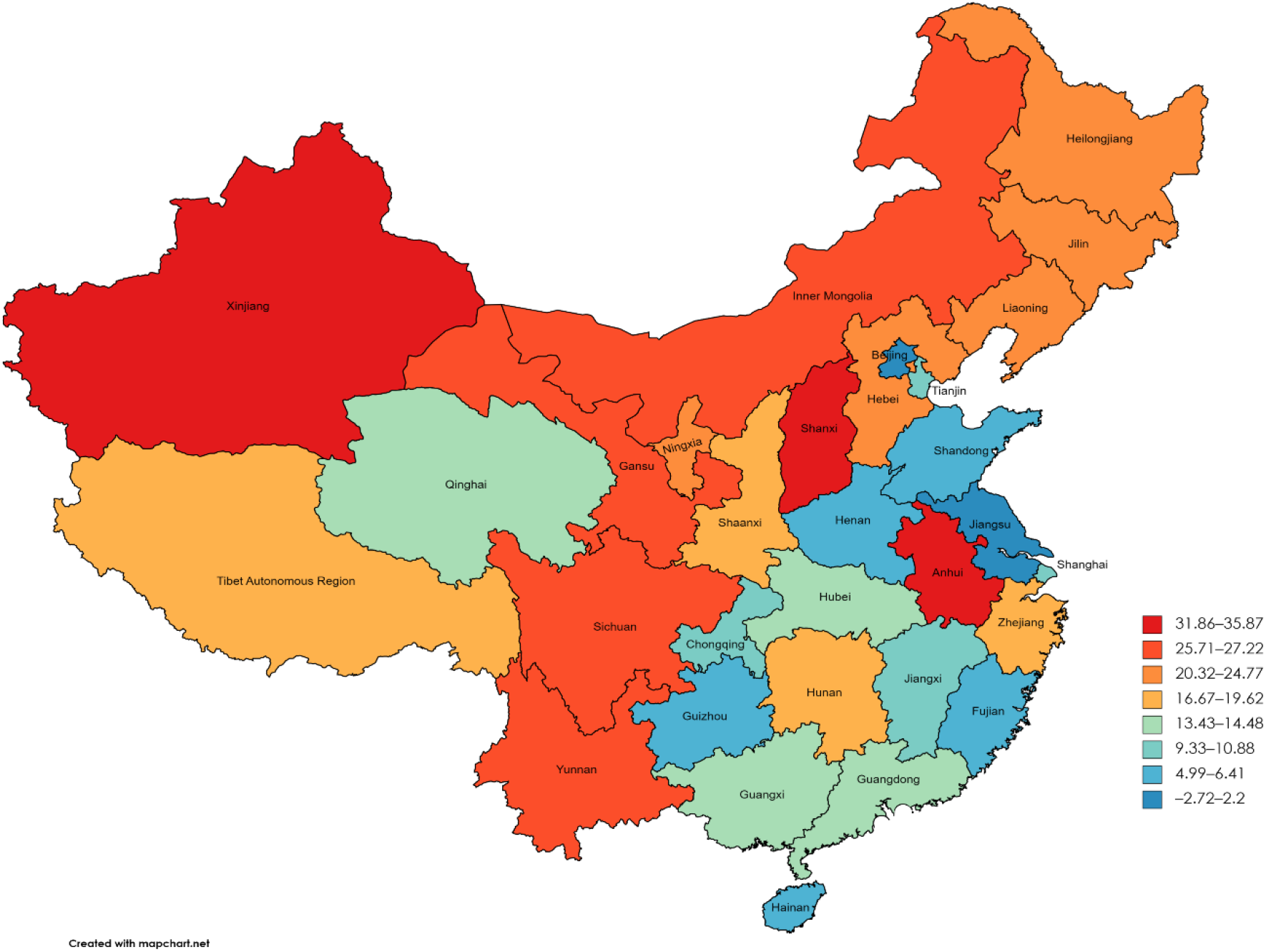
Distribution of Covid-19 excess death rates (%) in the 31 provinces, China, 2020–2022.

**Table 2.** Excess deaths and death rates in the four regions of China, 2020–2022.

| Region | Excess deaths in 1,000 |  |  |  | Excess death rate (%) |  |  |  | 2020–22<br>increase (%) |
| --- | --- | --- | --- | --- | --- | --- | --- | --- | --- |
|  | 2022 | 2021 | 2020 | Total | 2022 | 2021 | 2020 | 2020–22 |  |
| Eastern | 406 | 272 | 215 | 893 | 12.36 | 8.29 | 6.56 | 9.07 | 88.46 |
| The rest | 1231 | 1054 | 870 | 3154 | 22.44 | 19.23 | 15.86 | 19.18 | 41.49 |
| Central | 475 | 400 | 326 | 1202 | 19.96 | 16.79 | 13.71 | 16.82 | 45.85 |
| Western | 578 | 496 | 410 | 1484 | 23.86 | 20.46 | 16.91 | 20.41 | 41.13 |
| Northeast | 177 | 159 | 134 | 469 | 26.02 | 23.29 | 19.61 | 23.00 | 32.65 |
| Total | 1636 | 1327 | 1085 | 4048 | 18.67 | 15.13 | 12.38 | 15.40 | 50.81 |

**Table 3.** Results of independent samples t-tests comparing means of excess death rates between East and other regions of China, 2020–2022.

| Year | Region | N | Mean | Std.<br>Deviation | t | df | p | Cohen's<br>d |
| --- | --- | --- | --- | --- | --- | --- | --- | --- |
| 2020 | Eastern | 10 | 6.501 | 8.975 |  |  |  |  |
|  | The rest | 21 | 17.071 | 8.565 | 3.164 | 29 | 0.002 | 1.216 |
|  | Central | 6 | 16.001 | 11.777 | 1.828 | 14 | 0.045 | 0.944 |
|  | Western | 12 | 17.022 | 8.129 | 2.884 | 20 | 0.005 | 1.235 |
|  | Northeast | 3 | 19.406 | 3.419 | 2.377 | 11 | 0.018 | 1.565 |
| 2021 | Eastern | 10 | 8.157 | 7.146 |  |  |  |  |
|  | The rest | 21 | 20.484 | 8.328 | 4.021 | 29 | 0 | 1.545 |
|  | Central | 6 | 19.167 | 11.624 | 2.368 | 14 | 0.016 | 1.223 |
|  | Western | 12 | 20.5 | 7.797 | 3.838 | 20 | 0.001 | 1.643 |
|  | Northeast | 3 | 23.055 | 2.509 | 3.454 | 11 | 0.003 | 2.274 |
| 2022 | Eastern | 10 | 12.182 | 6.917 |  |  |  |  |
|  | The rest | 21 | 23.774 | 8.834 | 3.641 | 29 | 0.001 | 1.399 |
|  | Central | 6 | 22.863 | 12.361 | 2.239 | 14 | 0.021 | 1.156 |
|  | Western | 12 | 23.666 | 8.389 | 3.456 | 20 | 0.001 | 1.48 |
|  | Northeast | 3 | 26.026 | 1.073 | 3.352 | 11 | 0.003 | 2.207 |
| 2020–2022 | Eastern | 10 | 8.95 | 7.425 |  |  |  |  |
|  | The rest | 21 | 20.447 | 8.48 | 3.664 | 29 | 0 | 1.408 |
|  | Central | 6 | 19.344 | 11.837 | 2.177 | 14 | 0.024 | 1.124 |
|  | Western | 12 | 20.395 | 7.991 | 3.453 | 20 | 0.001 | 1.478 |
|  | Northeast | 3 | 22.861 | 2.29 | 3.114 | 11 | 0.005 | 2.05 |

**Table 4.** Results of independent samples t-tests comparing means of excess death rates of Chinese provinces with means of the OECD countries and the US states, 2020–2021.

| Region | N | Mean | Std. Deviation | t | df | p | Cohen's d |
| --- | --- | --- | --- | --- | --- | --- | --- |
| China | 31 | 15.085 | 9.73 |  |  |  |  |
| OECD | 35 | 13.583 | 11.339 | 0.574 | 64 | 0.284 | 0.141 |
| USA | 50 | 10.851 | 3.804 | 2.315 | 35.758 | 0.013 | 0.632 |

## Discussions

The findings indicate that China’s excess deaths were over 4 million by the end of 2022, while its excess death rate was 15.4%. These numbers are to be expected given China’s persisting inequalities, along with a large population and an upper-middle income economy. Inequalities have been blamed for worse Covid-19 outcomes in many parts of the world, including the US and the OECD (Covid Crisis Group, 2023; Bollyky et al., 2023; Berchet et al., 2023; OECD, 2023; Sachs et al., 2022; Lynch and Sachs, 2021). China’s rate was comparable to OECD countries. Furthermore, excess mortality estimation does not account for population aging or population increases. The Chinese population has been aging fast which may have led to an increase in death rates. That might have contributed slightly to the excess mortality. The study calculated the deaths in all provinces and found that death rates increased from 6.13 per 1,000 in 2015 to 6.35 in 2019. That was a 3.66% increase in five years or 0.73% each year. Total population of the 31 provinces increased from1,380 million in 2015 to 1,407 million in 2019, a 1.89% increase in five years. If the increases in death rates and population sizes had been taken into consideration, China’s excess deaths would have been a little bit lower. On the other hand, it should be noted that the US CDC does not report negative excess deaths, which are counted as zero excess deaths. Using the CDC’s methodology would have increased excess mortality in China by 50,750 deaths.

The striking disparities in excess mortality rates among the 31 provinces and the four regions should have also been expected. Such inequalities seem to follow China’s east-west divide. The more developed provinces in the eastern region displayed excess mortality at about the same rate as some more developed countries while disadvantaged western provinces had over twice the mortality rates. Such disparities were higher than those among the 50 US states. However, they were about the same among OECD countries, which include the world’s most developed countries as well as some developing countries.

It is difficult to figure out if and how provincial government Covid-19 control policy played a role in causing such inequalities in excess death rates, as China had a uniform policy under the central government. On the other hand, there were less disparities in excess mortality rates among the 50 US states, whose Covid-19 policy varied drastically. The main explainer is most likely to be pre-existing regional disparities in socio-economic development and sustainability.

The pandemic has further highlighted and intensified regional disparities in China. Continued efforts are required to address the underlying causes of regional inequality, including investment in infrastructure, education, healthcare, and promoting balanced regional development to ensure a more equitable and sustainable future for all regions in China. Further efforts need to be made to implement the UN 2030 Agenda and its SDGs, with attention to SDG 3 for improving health equity and SDG 10 for curtailing income inequalities (UN, 2015).

## Limitations

The study has several limitations and caution is advised when interpreting the findings. First, it should be noted that excess mortality data are only an estimation, as there is no way of knowing the exact number of deaths from 2020 to 2022 in China had the pandemic not happened. Second, there are different methods of estimation for excess mortality which may produce different results. Third, the study relied on Chinese government statistical publications, which have been subjected to questions regarding their reliability. GDP, income, and other economic achievements were often suspected to be over reported by local authorities. It is unknown if and how death rates or population sizes might have been manipulated. However, systematic under or over reporting of these figures would have little effect on the resulting excess mortality rates, which were based on percentage changes between time periods before and during the pandemic. Fourth, the NBS states that the provincial totals exclude population in the armed forces and are lower than the national totals of population (NBS, 2023a). When the total population is considered, China’s national excess deaths and death rate might be different. Finally, the study placed China in an international context. Such an approach may be problematic as China is so different from the US and the OECD.

## Conclusions

The study intends to contribute to the current debates over the true death toll of Covid-19 and possible lessons the world may learn from the pandemic. It reported an estimation of excess deaths in China. Striking regional differences were found in Covid-19 mortality among the 31 provinces and the four regions of China. Such regional disparities mirror the regional inequalities in socio-economic development that China has made significant efforts to reduce in the past decades. The indication was that China’s efforts in reducing regional inequalities did not achieve significant enough results to effectively control health disparities in the pandemic. Much work needs to be done to further reduce such inequalities to achieve a more sustainable society and prepare for the next pandemic or other major crises such as climate change.

## Data Availability

All data produced in the present work are contained in the manuscript

## Acknowledgements

The author thanks Tiffany Liu for editing the paper.

